# Deep learning screening of otoscopic images for ENT referral: development, validation and deployment of NHREF OtoScan

**DOI:** 10.64898/2026.11.04.26367286

**Authors:** L. G. Chandrasena, M. T. D. Lakshan, V. T. K. De Mel, B. Kumar Gautham

## Abstract

**Background:** Middle and external ear disease is common and largely treatable, but accurate otoscopic interpretation requires training that is scarce outside specialist ENT services. Published deep learning models for otoscopy report high accuracy, yet most are optimised for closed-set classification rather than the referral decision a non-specialist actually makes, report uncalibrated probabilities, and do not report the near-duplicate controls that video-derived otoscopic datasets demand. We developed, validated and deployed a screening aid framed around referral rather than diagnosis.

**Methods:** We merged 4,763 otoscopic images from 16 folders across three public datasets into 12 clinically meaningful classes. Every image was passed through a single field-of-view normalisation function (crop to the illuminated field, square, resize to 224 px, mask to one fixed circle) shared identically by training and inference, to remove the framing cue that otherwise identifies the source dataset. Because one source is video-derived, images were grouped by cosine similarity of generic ImageNet features (threshold 0.93, joined transitively) and whole groups assigned to a single split, giving 3,326 training, 719 validation and 718 test images. A MobileNetV2 backbone initialised from ImageNet was fine-tuned from block\_9\_expand. Probabilities were averaged over the eight dihedral transforms of each image and temperature-scaled. The referral score was one minus the probability of a normal ear, and its threshold was set on the validation abnormal ears as the highest value still meeting a target sensitivity of 97%, applied with an inclusive comparison. Model selection, calibration and threshold were frozen before the test split was scored once.

**Results:** On the 718 held-out test images, referral sensitivity was 97.0% (95% CI 95.1–98.2) and specificity 75.6% (95% CI 69.4–80.8), with AUROC 0.974 for abnormal versus normal; a cluster bootstrap over the 363 similarity groups in the test split widens those intervals to 93.8-99.6% and 66.0-83.4% respectively. Of 501 abnormal ears, 15 were not referred; 11 of these were myringosclerosis and none was cholesteatoma. Twelve-class macro F1 was 0.686, balanced accuracy 68.3%, and accuracy 85.0% (86.2% with chronic subtypes merged). Temperature scaling (T = 1.2) reduced expected calibration error from 0.0498 to 0.0430. A low-confidence flag fired on 4.0% of test images, separating 87.2% accuracy on unflagged from 31.0% on flagged cases. Recall was consistently highest on the largest source dataset and fell on the other two, for example acute otitis media 95.5% on source A versus 50.0% on source B.

**Conclusions:** A compact, calibrated model run on ordinary CPU hardware achieved the pre-specified 97% referral sensitivity with clinically interpretable failure modes. The source-stratified gap, and an earlier perceptual-hash split that inflated scores until it was corrected, indicate that headline accuracy in this field is strongly dependent on leakage control and acquisition style. The system is deployed with mandatory specialist review, a daily backtest and approval-gated retraining, so that prospective performance on local equipment is measured rather than assumed. It is research software and not a medical device.

**Registration:** Not applicable. This was retrospective model development on public data.

**Funding:** Funded by the Nawaloka Hospitals Research and Education Foundation.

## 1. Introduction

The decision a non-specialist makes after looking in an ear is usually not a diagnosis but a referral: does this ear need an ENT opinion? We built and deployed a model for that decision, and report here what it achieves and where it fails.

Disorders of the middle and external ear are among the most common reasons for primary care consultation worldwide, and most are treatable when identified. Chronic suppurative otitis media and cholesteatoma carry a risk of permanent hearing loss and intracranial complication when they go unrecognised, while conditions such as cerumen impaction and myringosclerosis need little or no intervention. Distinguishing between these on otoscopy is a learned skill, and in settings where ENT specialists are few, the clinician holding the otoscope is frequently not the clinician best placed to interpret what they see.

Deep learning on otoscopic images has been studied for several years, and reported accuracies are often high (Cha et al., 2019; Başaran et al., 2020). Three gaps recur in that literature and motivated the present work.

First, most published models are framed as closed-set classifiers over a fixed condition list, and are evaluated with accuracy or macro F1. Neither quantity describes the error that matters clinically. A model that is right about which disease is present, but occasionally confident that a diseased ear is normal, is more dangerous than one that is vague about the disease but reliably flags abnormality. We therefore make the binary refer/do-not-refer decision the primary endpoint, fix its operating point in advance on a validation split, and report the differential diagnosis as secondary.

Second, reported probabilities are rarely calibrated. A clinician reading “82% confident” reasonably expects that to mean something about frequency. Without calibration it does not, and a screening aid whose confidence cannot be interpreted is difficult to use safely alongside clinical judgement.

Third, and most consequential for how published numbers should be read, several public otoscopic datasets are derived from video. The same ear then appears across long runs of frames. A random split, or a split grouped only by perceptual hash, places near-identical views of one ear on both sides of the partition and inflates the reported score. We encountered this directly: an earlier version of our own pipeline used hash-based grouping and left roughly half of one source’s test images with a near twin in training. The figures reported here come from the corrected split, and the difference between the two is large enough that we treat it as a finding rather than a footnote.

A fourth consideration is what happens after development. A model validated once on retrospective public data says little about performance on a particular clinic’s otoscopes, operators and patient mix. We therefore treat deployment as the beginning of measurement rather than its conclusion: every result is reviewed by a surgeon, those verdicts accumulate as a prospective record of local performance, a scheduled backtest re-runs the model in service against them, and retraining is gated on both a frozen test set and an explicit human approval.

This paper reports the development and retrospective validation of NHREF OtoScan, a screening aid that recommends ENT referral from a single otoscopic image, names the most likely of twelve conditions with a calibrated differential, and displays the image regions driving the finding. We describe the leakage and confounding controls in enough detail to be reproduced, report performance stratified by source dataset, and describe the deployed system and its review loop. We make no claim of prospective clinical validation; the system is research software, is not a medical device, and is used only with mandatory specialist review.

## 2. Materials and Methods

### 2.1 Data sources and class definitions

We assembled 4,763 otoscopic images of the tympanic membrane and external canal from 16 labelled folders, drawn from openly licensed otoscopic image datasets published on Kaggle and referred to throughout as sources A (3,014 images), B (956) and C (793). Source attribution is recoverable from each dataset’s filename convention and is recorded for every image in the study manifest, which is distributed with the code.

All images were obtained from three openly available otoscopic image datasets published on Kaggle (ucimachinelearning, 2025; Başaran, 2022; Zaka ur Rehman, 2026), summarised in Table 1. We downloaded these collections and assigned every image to one of the 12 classes used in this study ourselves, because the source collections use inconsistent and partly overlapping condition vocabularies. The source labels A, B and C used throughout this paper refer to these three collections, and every image’s source is recorded in the manifest distributed with the code.

**Table 1.** Source datasets.

| **Source** | **Dataset** | **Depositor** | **Images** | **Folders** | **Licence** |
|---|---|---|---|---|---|
| A | Oto-Endoscopic Image Dataset | ucimachinelearning | 3,014 | 5 | CC0 1.0 (public domain) |
| B | Tympanic membrane / eardrum dataset / otitis media | ebasaran | 956 | 9 | Not stated |
| C | OtitisMediaKPJ: Otoscopic Dataset for Otitis Media | zakaurrehmanmmu | 793 | 6 | CC BY-NC-SA 4.0 |
|  | **Total** |  | **4,763** | **20** |  |

The three differ in provenance and in how they were assembled, which is why they are analysed separately in the Results. Source A is a set of oto-endoscopic images distributed in five balanced class folders. Source B comprises otoscope images each assessed by three ENT specialists, with low-quality images excluded by its depositors; its dataset record asks that Başaran et al. (2020) be cited alongside it. Source C comprises de-identified images collected during routine ENT consultations at KPJ Healthcare Specialist Hospitals using a digital video otoscope, each independently reviewed and annotated by ENT specialists.

Two discrepancies in the source records are worth stating. Source A’s dataset card describes itself as balanced with 614 images per category while listing per-class counts of 601 to 605; our own counts, given in Table 3, are taken directly from the files and are the figures used throughout. The 20 source folders in Table 1 reduce to the 16 distinct folder names reported below because four condition names recur across datasets.

The 16 source folders use inconsistent vocabulary, and two pairs name the same entity: Ear wax with Cerumen_Impaction, and tympanoskleros with Myringosclerosis. Three folders (foreign body, pseudomembrane and ventilation tube) contained fewer than 20 images each and were pooled into a single *Other Findings* class rather than modelled separately. The resulting 12 classes are defined in Table 2.

**Table 2.** The 12 study classes and their clinical definitions.

| **Class** | **Clinical definition** |
|---|---|
| Acute otitis media | Acute middle ear infection; bulging, erythematous tympanic membrane |
| Otitis media with effusion | Fluid behind an intact tympanic membrane without acute infection |
| Chronic otitis media (unspecified) | Chronic middle ear disease; subtype not recorded in the source data |
| Chronic suppurative otitis media | Persistent perforation with chronic discharge |
| Chronic OM, active, no cholesteatoma | Perforation with active discharge, no cholesteatoma |
| Chronic OM, inactive | Dry tympanic membrane perforation without discharge |
| Chronic OM with cholesteatoma | Keratinising squamous epithelium in the middle ear; erosive |
| Myringosclerosis | Calcified white plaques in the tympanic membrane; includes tympanosclerosis |
| Cerumen impaction | Ear wax occluding the canal or obscuring the tympanic membrane |
| Otitis externa | Inflammation of the external ear canal |
| Other findings | Foreign body, pseudomembrane or ventilation tube, pooled |
| Normal ear | No abnormality detected |

**Table 3.** Images per class and split.

| **Class** | **Train** | **Validation** | **Test** | **Total** |
|---|---|---|---|---|
| Normal ear | 1,013 | 217 | 217 | 1,447 |
| Acute otitis media | 572 | 123 | 123 | 818 |
| Cerumen impaction | 518 | 112 | 112 | 742 |
| Myringosclerosis | 443 | 95 | 95 | 633 |
| Chronic OM (unspecified) | 420 | 91 | 91 | 602 |
| Otitis media with effusion | 95 | 21 | 21 | 137 |
| Chronic OM, inactive | 71 | 16 | 16 | 103 |
| Chronic OM, active, no cholesteatoma | 59 | 13 | 13 | 85 |
| Chronic suppurative OM | 43 | 10 | 10 | 63 |
| Chronic OM with cholesteatoma | 43 | 10 | 9 | 62 |
| Otitis externa | 29 | 6 | 6 | 41 |
| Other findings | 20 | 5 | 5 | 30 |
| **Total** | **3,326** | **719** | **718** | **4,763** |

Five of the twelve classes are chronic otitis media subtypes. In the source data, *Chronic otitis media (unspecified)* functions as a parent label of the other four rather than a disjoint category, so subtype-level predictions should be read with that overlap in mind. We therefore report a coarse accuracy with the five chronic subtypes merged, and the deployed interface presents the summed chronic probability alongside the individual subtypes, since the group total is more reliable than any single member.

One absence is worth stating explicitly, because the project was originally scoped around it: none of the three sources contains otomycosis images, so fungal otitis externa is not a recognised class and cannot be detected. The deployed system allows a reviewing surgeon to record it as a free-text diagnosis, which is the mechanism by which enough local cases could be accumulated to add the class later.

All images are retrospective, de-identified and were already publicly available before this study. No patient-identifying information was available to, or required by, the investigators. Because the class assignment was performed by the authors rather than inherited from the source collections, the labels carry our interpretation of each image’s original folder name, and the taxonomy mapping we applied is distributed with the code so that it can be inspected or revised.

### 2.2 Field-of-view normalisation

The three source datasets frame the ear differently: one applies a hard black circular mask, one fills the frame edge to edge, and one sits between the two. Because 8 of the 12 classes are drawn from a single source, that framing is close to a label. Before normalisation we measured 99% of images from one source as having pure black corners against 17% from another, a cue sufficient for a network to score well by identifying the dataset rather than reading the ear.

Every image therefore passes through one normalisation function, in four steps:

1. Locate the illuminated field of view and crop to it. A pixel counts as illuminated above a luminance of 20, chosen because the darkest source retains roughly that value outside its circular mask.
2. Centre-crop to a square.
3. Resize to the 224 × 224 px network input.
4. Set every pixel outside one fixed inscribed circle to black, at a radius of 0.95 of the half-width. That radius was chosen from the measured radial luminance profile of each source: all three keep their content inside 0.95, while the harder mask edges of two of them fall between 0.95 and 1.0.

After step 4 the region outside the circle is identical for every image from every source and can no longer carry information about provenance.

This function is implemented once, in a single module, and imported by both the training pipeline and the inference service. That is a deliberate design constraint rather than an implementation detail: a preprocessing pipeline reimplemented on the serving side is a standing opportunity for training-serving skew, and here the two cannot drift apart because there is only one definition. The training pipeline caches the normalised copy of each image, so the network sees bytes produced by the same code path that will run in the clinic.

### 2.3 Partitioning and near-duplicate control

Near-duplicate images are not spread evenly across the three sources, and the difference is stark. In source A the same ear recurs across long runs of near-identical frames, consistent with extraction from oto-endoscopic video: 672 of its 1,040 similarity groups hold more than one image, and its largest holds 36. Sources B and C are predominantly singletons, with largest groups of 3 and 6 images respectively. Any split that treats images as independent will therefore place near-identical views of the same source A ear on both sides of the partition, and the resulting test score measures memorisation rather than generalisation.

Our first implementation grouped images by perceptual hash. This proved inadequate. A perceptual hash catches only frames that are near-pixel-identical; as the otoscope moves, successive frames of the same ear drift past the hash threshold while remaining obviously the same ear to a human observer. Auditing that split, roughly half of source A’s test images had a near twin in training, and the scores for that source were correspondingly inflated. We report this because the failure is invisible in the headline metric, since the model simply looks better, and because the same hazard applies to any otoscopic dataset built from video.

The corrected procedure groups by semantic rather than pixel similarity:

5. Each normalised image is embedded using generic ImageNet MobileNetV2 features. These are pretrained features, not features learned from this dataset, so grouping cannot be contaminated by the labels.
6. Any two images of the same class with cosine similarity ≥ 0.93 are joined, and groups are formed as the transitive closure (connected components) of that relation.
7. Whole groups are assigned to exactly one split, so every frame of a given ear stays together.

Two checks constrain the threshold. At 0.93, no group exceeds one tenth of its class, which bounds the risk that transitive joining has chained genuinely distinct ears into one component. After splitting, the maximum similarity between any test image and any same-class training image was 0.92999, confirming the constraint holds in the delivered partition. Because that grouping compares images only within a class, a further review was carried out across classes: every test and validation image was compared with every training image irrespective of label, using the same features and the same 0.93 threshold. Its results are given in section 3.2.

Images were then split 70/15/15 by group under a fixed random seed (1337), giving 3,326 training, 719 validation and 718 test images. The validation split carried every decision that required looking at data: architecture and fine-tuning choices, early stopping, the calibration temperature, and the referral threshold. The test split was scored once, after all of those were frozen. No test image contributed to any choice reported in this paper.

The complete manifest, which records for every image its path, source folder, source dataset, class, group identifier and split, is distributed with the code so that the partition can be audited or reproduced exactly.

### 2.4 Model architecture and training

We used MobileNetV2 (Sandler et al., 2018) initialised from ImageNet weights, with a global average pooling layer, dropout at 0.3, and a single softmax dense layer over the 12 classes. The architecture was chosen for deployability: the target installation is an ordinary clinic computer with no GPU, and the pooled-linear head also yields exact class activation maps, described below.

Training ran in two stages. The backbone was first frozen and the head trained for 10 epochs with Adam at a learning rate of 1×10⁻³. The backbone was then unfrozen from the block_9_expand layer onward and fine-tuned for a further 18 epochs under a decaying learning-rate schedule. Batch size was 16, constrained by the four-core CPU and available memory of the development machine. Class imbalance was handled with square-root inverse-frequency class weights. Checkpoint selection used validation macro F1; the best epoch was 26, with validation macro F1 of 0.716.

Augmentation was chosen to match the invariances of the clinical setting rather than applied generically:

- **Rotation through any angle, and horizontal flip.** Otoscope orientation is arbitrary, so every orientation is label-preserving. The fixed circular field of view is invariant under rotation, so no information from outside the circle can leak in.
- **Zoom in only, up to 12%.** Zooming out would draw a black ring inside the circle, a pattern the service never produces at inference.
- **Brightness (±20 levels) and contrast (0.85–1.15).** Illumination varies widely between devices and operators.
- **Saturation 0.9–1.1 and hue ±0.02.** Colour is diagnostic in this domain (erythema, discharge, keratin debris), so chromatic augmentation is deliberately narrow; aggressive hue shifts would destroy signal rather than add robustness.

After augmentation the fixed circular mask is reapplied, so the network always sees exactly the field of view the service will present. Training took approximately 40 minutes on four CPU cores.

### 2.5 Test-time augmentation, calibration and the referral threshold

#### Orientation averaging

At inference, logits are averaged over all eight dihedral transforms of the image (four rotations, each with and without reflection) before the softmax. The clinical justification is that the result should not depend on how the otoscope happened to be held; the empirical justification is that validation macro F1 rose from 0.716 for a single view to 0.733 with averaging. The property is asserted in the automated test suite, so a future change that breaks orientation invariance fails the build rather than degrading silently in service.

#### Temperature scaling

Following Guo et al. (2017), a single scalar temperature was fitted on the validation split alone, by grid search minimising negative log likelihood, giving T = 1.2. Temperature scaling preserves the ranking of classes and therefore cannot change which condition is named; it changes only how confident the stated probability is.

#### The referral score

The probability that an ear is abnormal is defined as one minus the calibrated probability of the normal class. This is deliberately not a separately trained binary head. Deriving the screening decision from the same distribution that produces the differential guarantees the two can never disagree: a system cannot recommend no referral while naming cholesteatoma as the leading finding.

#### Threshold selection

The referral threshold was chosen on the validation abnormal ears as the highest threshold whose sensitivity still met the 97% target, which maximises specificity subject to meeting that target, giving 0.2320. In implementation the abnormality scores of the validation abnormal ears are sorted in ascending order, at most floor((1 - 0.97)n) of them are permitted to fall below the threshold, and the threshold is set at the next score above that point. The comparison is inclusive: an ear whose abnormality score equals the threshold is referred. Where scores tie at the threshold value the realised sensitivity therefore meets or exceeds the target rather than falling below it, so ties resolve toward referral. The 97% target was set before the test split was scored, as a clinical policy decision: in screening for conditions that include cholesteatoma, a missed abnormality is materially worse than an unnecessary specialist opinion. Thresholds for four alternative targets were computed the same way on validation and are reported in Results as a sensitivity analysis; the deployed value remains the 97% point, and changing it is a one-line configuration change requiring no retraining.

#### Low-confidence flag

A finding is marked low-confidence when the margin between the top two classes is below 0.15, or when the entropy of the distribution exceeds 80% of its theoretical maximum. This flag qualifies the named condition only; it never suppresses or alters the referral recommendation.

The resulting behaviour is asymmetric by design. Where the model is uncertain whether an ear is normal, the decision resolves toward referral, and the interface states that it has done so.

### 2.6 Explanation of findings

The network ends in global average pooling followed by a single linear layer, the arrangement that yields class activation maps (Zhou et al., 2016). Under it the class activation map is not an approximation of the model’s reasoning but an algebraic identity: the logit for a class is exactly the spatially pooled inner product of the final feature map with that class’s weight vector. Projecting the feature map onto the same weight vector therefore recovers, per spatial location, that location’s additive contribution to the logit.

This matters practically. Gradient-based attribution methods estimate what a different architecture might have attended to, and can disagree with the prediction they purport to explain. Here the explanation and the prediction come from one forward pass and cannot diverge. The map is computed from the unrotated view even when orientation averaging is active, so the highlighted regions align with the image the clinician is shown.

The map is rectified at zero, normalised to its peak, and rendered as a blue tint over the normalised image. The interface presents it beside the image as the model received it, so a clinician can see both what the model looked at and what it was given to look at, the latter being how field-of-view problems, such as an image dominated by canal wall, become visible.

### 2.7 Deployed system and clinical governance

The model is deployed as a web service. A surgeon signs in, enters the patient’s hospital record number, age, sex and the ear examined, then uploads an otoscopic image or captures one directly from a USB video otoscope or endoscope. Analysis takes approximately one third of a second on CPU. The result is written to the patient record with timestamp, clinician and the model version that produced it.

#### Mandatory review

Every result is reviewed. The surgeon either confirms the finding, selects the correct condition from the list, or records a diagnosis outside the twelve classes as free text. Reviews are append-only. This serves two purposes: no automated output stands as a clinical record without specialist endorsement, and the accumulated verdicts constitute a prospective measurement of performance on this clinic’s own patients and equipment, which retrospective public data cannot provide.

#### Daily backtest

Once a day the model in service is re-run over every reviewed case and compared against the surgeons’ verdicts. Cases that a model was retrained on, and copies of original dataset images, are excluded so that no model is ever scored on its own training data. The dashboard raises an alert when recent referral sensitivity falls below 90%, or when recent agreement falls more than 10 percentage points below earlier agreement, subject to a minimum case count. The backtest runs in a separate process and never modifies the model.

#### Approval-gated retraining

Retraining may be started by an administrator once at least 50 reviewed cases exist, and follows a fixed protocol. Approximately 30% of reviewed *patients* form a reserved evaluation cohort that is excluded from training and calibration. Membership is fixed by a hash of the patient’s identifier rather than drawn afresh at each run, so the same patients are reserved every time and a patient reserved once is never later trained on. Splitting by patient also keeps both ears and repeat visits for one patient on a single side of the split. Any uploaded image identical to an image in the frozen validation or test set is excluded from training, so that the comparison cannot be contaminated. The candidate is calibrated and evaluated exactly as the incumbent was, then both are scored on the held-out local patients. A candidate that is worse than the incumbent on either the frozen test set or the local patients cannot be approved. A candidate that passes still requires explicit administrator approval, which switches the active model without a restart and regenerates the validation report.

We chose supervised, gated retraining over continuous online learning deliberately. A model that updated itself from every clinician click could be degraded by a small number of mistaken reviews with no one noticing, and in a clinical setting silent degradation is the failure mode that matters most.

#### Privacy and auditability

Patients are identified by hospital record number only; names, dates of birth and contact details are never stored. Images are held outside the web root under random names, because clinic filenames frequently contain patient names, and uploaded filenames are never logged or retained. Stored images are capped at 200 KB, re-encoded only when larger, with the analysed image being the stored image so that record, backtest and retraining all reference identical bytes. Sign-in, examination, review, account change, deletion and model approval are each written to an append-only audit log. The service runs as a dedicated database role owning only its own database.

At the time of writing the system is deployed and operational but has not yet accumulated the reviewed cases required for a first retraining run, and no prospective performance figures are therefore reported in this paper.

### 2.8 Statistical analysis

The primary endpoint was referral sensitivity and specificity on the held-out test split at the pre-specified threshold, with abnormal defined as any of the eleven non-normal classes. Binomial proportions are reported with 95% Wilson score intervals (Wilson, 1927), which behave better than the normal approximation at the proportions near 1 that arise here.

Discrimination independent of threshold is summarised by AUROC for abnormal versus normal.

Secondary endpoints were twelve-class macro F1, balanced accuracy, overall accuracy, coarse accuracy with the five chronic subtypes merged, and per-class precision, recall and F1 with support. Macro-averaging weights every class equally and is therefore the more demanding summary given the class imbalance; we report accuracy alongside it for comparability with the literature, not as the headline figure.

Calibration was assessed by expected calibration error over equal-width confidence bins and by negative log likelihood, each computed before and after temperature scaling.

Performance was additionally stratified by source dataset, restricted to the four conditions appearing in more than one source. This is the analysis most directly relevant to generalisation: where a condition exists in several sources, consistent recall across them indicates the model is reading the ear rather than the acquisition style.

#### Group-aware intervals

Because near-duplicate frames make images within a similarity group statistically dependent, referral sensitivity and specificity were also given group-aware intervals by a cluster bootstrap: the similarity groups of the test split were resampled with replacement 2,000 times under a fixed seed, and the 2.5th and 97.5th percentiles of the resulting distributions taken as the interval.

All reported figures were produced by a single scripted evaluation run over the frozen test split, which also regenerates the validation report distributed with the code. No figure in this paper was computed by hand or transcribed, and the script can be re-run to reproduce every value.

Analyses were performed in Python 3.11 with TensorFlow 2.15 and NumPy. No formal sample size calculation was performed, as the study used the entirety of three fixed public datasets; the resulting precision is reported as confidence intervals throughout, and is explicitly poor for the smallest classes.

## 3. Results

### 3.1 Dataset composition

The 4,763 images divided into 3,326 training, 719 validation and 718 test images. Class support is markedly uneven: the three largest classes (normal ear, acute otitis media and cerumen impaction) account for 3,007 of 4,763 images (63.1%), while six classes contribute fewer than 110 images each across all splits.

By source, 3,014 images came from source A, 956 from source B and 793 from source C. Eight of the twelve classes are drawn from a single source, which is the central threat to validity in this corpus and the reason for both the field-of-view normalisation and the source-stratified analysis reported below.

In the test split, 501 of 718 images (69.8%) were abnormal and 217 (30.2%) were normal.

### 3.2 Screening performance

At the pre-specified threshold of 0.2320, the model referred 97.0% of abnormal ears and correctly withheld referral for 75.6% of normal ears. The target sensitivity of 97%, fixed on validation before the test split was scored, was met almost exactly on test (97.0%), indicating the operating point transferred without recalibration.

The confusion at that operating point was 486 true positives, 15 false negatives, 164 true negatives and 53 false positives.

**The 15 missed abnormal ears** are the result that matters most for safety, and their composition is reassuring. Eleven were myringosclerosis, a calcified plaque of the tympanic membrane that is usually benign and often an incidental finding. Two were acute otitis media, one cerumen impaction, and one fell in the pooled *Other findings* class. **No cholesteatoma was missed**, and no chronic suppurative otitis media was missed. These are the two conditions in this class list where delayed recognition carries the clearest risk of permanent harm.

The 53 false positives represent normal ears sent for an ENT opinion they did not need. At a prevalence of 69.8% abnormal in this test set the resulting PPV is 90.2%, but that figure will fall in a screening population with lower disease prevalence, and should not be carried over to such a setting unchanged.

An AUROC of 0.974 indicates that discrimination between abnormal and normal is strong independently of where the threshold is placed, so the specificity cost reported here is a consequence of the deliberately aggressive operating point rather than of weak underlying separation.

#### Group-aware uncertainty

The 718 test images fall into 363 similarity groups, so observations within a group are not independent and the Wilson intervals in Table 4, which assume independence, understate uncertainty. A cluster bootstrap over those groups, resampling them with replacement 2,000 times, gives a 95% interval of 93.8 to 99.6% for referral sensitivity and 66.0 to 83.4% for specificity, against the Wilson intervals of 95.1 to 98.2% and 69.4 to 80.8%. The group-aware intervals are the wider and the more appropriate of the two, and the figures in Table 4 should be read with that in mind.

**Table 4.** Primary screening endpoint, 718 held-out test images.

| **Measure** | **Result** | **95% CI** |
|---|---|---|
| Referral sensitivity (abnormal ears referred) | 97.0% | 95.1 to 98.2% |
| Referral specificity (normal ears not referred) | 75.6% | 69.4 to 80.8% |
| Positive predictive value | 90.2% |  |
| Negative predictive value | 91.6% |  |
| AUROC, abnormal vs normal | 0.974 |  |
| Referral threshold | 0.2320 | fixed on validation |

**Table 5.** Composition of the 15 non-referred abnormal ears.

| **Condition** | **Missed** |
|---|---|
| Myringosclerosis | 11 |
| Acute otitis media | 2 |
| Cerumen impaction | 1 |
| Other findings | 1 |
| Cholesteatoma, suppurative OM, all other classes | 0 |

**Table 6.** Per-class performance on the test split.

| **Class** | **Precision** | **Recall** | **F1** | **Support** |
|---|---|---|---|---|
| Chronic OM (unspecified) | 1.000 | 1.000 | 1.000 | 91 |
| Cerumen impaction | 0.982 | 0.955 | 0.968 | 112 |
| Acute otitis media | 0.836 | 0.870 | 0.853 | 123 |
| Normal ear | 0.774 | 0.899 | 0.832 | 217 |
| Chronic OM, active, no cholesteatoma | 0.667 | 0.923 | 0.774 | 13 |
| Myringosclerosis | 1.000 | 0.632 | 0.774 | 95 |
| Chronic OM with cholesteatoma | 0.700 | 0.778 | 0.737 | 9 |
| Otitis media with effusion | 0.630 | 0.810 | 0.708 | 21 |
| Chronic OM, inactive | 0.818 | 0.562 | 0.667 | 16 |
| Other findings | 0.400 | 0.400 | 0.400 | 5 |
| Chronic suppurative OM | 0.400 | 0.200 | 0.267 | 10 |
| Otitis externa | 0.500 | 0.167 | 0.250 | 6 |
| **Macro average** |  |  | **0.686** | **718** |

#### Cross-class near-duplicate review

The grouping procedure compares images only within a class, so a separate review compared every test image with every training image irrespective of label. Eleven of the 718 test images (1.5%) had a training image of a different class at or above the 0.93 similarity threshold, and three of those pairs were effectively identical at a cosine similarity of 1.000. Seven pairs lay within source B and four between sources A and B, and they concentrate on four training files: one image labelled normal matched five test images labelled acute otitis media, one labelled acute otitis media matched three test images labelled normal, one matched two test images labelled otitis externa, and one labelled other findings matched a test image labelled chronic suppurative otitis media. These are duplicate or near-duplicate images carrying inconsistent labels rather than further views of one ear. Excluding all eleven moves referral sensitivity by less than a tenth of a percentage point, from 97.01% to 96.96%, and raises specificity from 75.58% to 76.64%, so the reported figures are not materially affected. The direction is what should be expected: a near-twin in training that carries a different label teaches the wrong answer for that image rather than inflating its score.

### 3.3 Twelve-class classification

Twelve-class macro F1 was 0.686, balanced accuracy 68.3%, and overall accuracy 85.0%. Merging the five chronic otitis media subtypes into a single chronic class raised accuracy to 86.2%, confirming that a meaningful share of the residual error is subtype confusion within chronic disease rather than failure to recognise chronic disease.

Four observations follow from this table, and the first two are warnings rather than achievements.

#### The perfect score should be distrusted

Chronic otitis media (unspecified) achieved precision and recall of 1.000 on 91 test images. This class is drawn from a single source dataset. A perfect score on a single-source class is more plausibly a signature of that source’s acquisition style than evidence of diagnostic skill, and it should not be quoted as such.

#### Myringosclerosis shows a characteristic asymmetry

Precision is 1.000 but recall only 0.632: when the model names myringosclerosis it is always right, but it misses more than a third of cases. This single behaviour accounts for 11 of the 15 referral misses. Clinically the consequence is mild, since myringosclerosis is usually benign, but it is the dominant failure mode of the system and is stated here plainly.

#### The smallest classes are not usable as reported

Otitis externa (recall 0.167 on 6 images), chronic suppurative OM (0.200 on 10) and *Other findings* (0.400 on 5) have support too small for the point estimates to be meaningful. These figures establish only that the model does not reliably recognise these conditions; they do not establish how badly.

#### Normal ear recall is high and precision moderate

(0.899 / 0.774), which is the expected signature of a system tuned to over-refer: the model is reluctant to assign normal, so when it does so it is usually correct, and some abnormal ears are pulled into the normal class, which is exactly the error the referral threshold is set to intercept.

### 3.4 Calibration and choice of operating point

#### Calibration

The fitted temperature was 1.2, indicating the uncalibrated network was mildly overconfident. On the test split, expected calibration error fell from 0.0498 to 0.0430 and negative log likelihood from 0.391 to 0.380. The improvement is modest because the model was not badly miscalibrated to begin with; the value of the step is that a stated confidence can now be read as an approximate frequency, which is the premise on which a clinician can weigh the model’s output against their own impression.

#### Low-confidence flag

The flag fired on 4.0% of test images. Accuracy was 87.2% on unflagged cases against 31.0% on flagged ones. The flag therefore does useful work: it identifies a small subset where the named condition is close to unreliable, and it does so without touching the referral recommendation.

#### Operating points

The referral threshold encodes a clinical policy, not a technical optimum. Table 7 reports four alternatives, each with its threshold chosen on the validation split by the same procedure and then scored on test, so the comparison is honest rather than a post-hoc sweep of the test set.

**Table 7.** Referral threshold sensitivity analysis. Thresholds chosen on validation; performance measured on the 718-image test split (501 abnormal, 217 normal).

| **Target sensitivity** | **Threshold** | **Test sensitivity** | **Test specificity** | **Abnormal ears missed** | **Unnecessary referrals** |
|---|---|---|---|---|---|
| 90% | 0.4849 | 90.0% | 87.1% | 50 | 28 |
| 93% | 0.3812 | 93.4% | 83.4% | 33 | 36 |
| 95% | 0.2857 | 95.0% | 78.3% | 25 | 47 |
| **97% (deployed)** | **0.2320** | **97.0%** | **75.6%** | **15** | **53** |
| 99% | 0.1891 | 99.0% | 72.8% | 5 | 59 |

The trade is steep at the top of the range. Moving from 90% to 97% target sensitivity prevents 35 missed abnormal ears at a cost of 25 additional unnecessary referrals; moving from 97% to 99% prevents a further 10 misses for 6 more unnecessary referrals. We selected 97% as the deployed policy because the conditions at stake include cholesteatoma, but the choice is a clinical judgement for the supervising ENT service rather than a property of the model, and it is a single configuration value requiring no retraining to change. A related clinical question, whether uncomplicated cerumen impaction should trigger referral at all rather than local management, is likewise a policy decision outside the scope of this evaluation.

### 3.5 Performance by source dataset

Four conditions appear in more than one source dataset, allowing recall to be compared across acquisition styles while holding the condition fixed. This is the most informative generalisation evidence the corpus supports, and it is the least flattering result in the paper.

Recall is highest on source A for every condition, and source A is also the largest contributor to training (3,014 of 4,763 images). The gap is substantial: acute otitis media is recognised in 95.5% of source A images against 50.0% of source B images, and normal ears in 100% against 81.7%. Field-of-view normalisation removed the framing cue, and the near-duplicate grouping removed same-ear leakage, but differences in camera hardware, illumination, focus and compression evidently remain and the model remains sensitive to them.

The practical reading is that the pooled headline figures in Table 4 are weighted toward the acquisition style the model saw most, and that the source B and C columns are the better estimate of what to expect from unfamiliar equipment. A deploying site should assume performance closer to the lower columns until it has measured its own.

#### Input characteristics

For reference when assessing whether a new site’s images resemble the training distribution, the corpus had median brightness 136 (1st–99th percentile 69–188) and median Laplacian sharpness 17.2 (1st–99th percentile 3.0–564). The wide sharpness range reflects genuinely variable focus quality across sources.

#### Runtime

Inference, including the eight-view orientation averaging, takes approximately one third of a second per image on an ordinary CPU, with the model occupying about 280 MB in memory. No GPU is required at training or inference, which is what makes deployment on routine clinic hardware practical.

## 4. Discussion

### Principal findings

A compact model running on ordinary CPU hardware met a pre-specified 97% referral sensitivity on a held-out test split, with 75.6% specificity and AUROC 0.974. The failure modes are clinically legible: of 15 missed abnormal ears, 11 were myringosclerosis, a usually benign finding, and no cholesteatoma or chronic suppurative otitis media was missed. Twelve-class macro F1 of 0.686 is modest, and we regard it as the honest figure for this corpus rather than a disappointment, since it reflects six classes with fewer than 20 test images and five overlapping chronic subtypes.

### On comparison with published accuracies

Reported accuracies for otoscopic deep learning are frequently higher than ours, and we would caution against reading that difference as a difference in model quality. Our own pipeline produced materially better numbers before its near-duplicate control was corrected. Because public otoscopic datasets include video-derived material, and because perceptual hashing does not catch frames that drift as the otoscope moves, a split can appear sound while leaving half of one source’s test images with a near twin in training. We can speak only to our own case, but the lesson generalises: in this domain the reported score is a function of the partitioning method, and papers that do not state how near-duplicates were handled are difficult to compare against. We report the grouping method, the similarity threshold, the maximum test-to-train similarity achieved (0.92999) and the full manifest, so that ours can be checked.

### Source dependence is the main scientific caveat

The gap between 95.5% and 50.0% recall for acute otitis media across two sources is larger than the gap between most published models. Field-of-view normalisation was effective against the framing confound, having been introduced precisely because 99% of one source had black corners against 17% of another, but it cannot neutralise differences in sensor, optics, illumination and compression. Any single-centre validation of an otoscopic model, including a future one of ours, should be read with this in mind.

### Design choices framed by clinical use

Three decisions follow from treating the referral decision as primary. Deriving the abnormality score as one minus the calibrated probability of normal, rather than training a separate binary head, makes it structurally impossible for the screening recommendation to contradict the differential. Fixing the operating point on validation to a sensitivity target, rather than reporting the best achievable accuracy, means the deployed behaviour was specified as policy before it was measured. And resolving uncertainty toward referral, while saying so in the interface, matches the asymmetry of harms in this setting.

The explanation design follows the same logic. Because the network ends in average pooling and one linear layer, the activation map is an identity rather than an approximation, so the clinician is shown where the evidence actually came from and not an estimate of where it might have. Displaying the normalised image alongside it also surfaces preprocessing failures, which would otherwise be invisible.

### Deployment as measurement

The most defensible claim available from retrospective public data is a narrow one, and we have tried not to exceed it. What the deployment adds is not performance but measurement: mandatory specialist review turns routine use into a prospective record of local accuracy, the daily backtest detects degradation against those verdicts, and retraining is gated on both a frozen test set and explicit human approval. We chose this over continuous learning because a model that updates itself from every click can be degraded by a handful of mistaken reviews without anyone noticing, and silent degradation is the failure mode least likely to be caught in clinical use.

### Comparison with clinical practice

We did not compare the model against clinicians, and the comparison the system is designed for is not model-versus-expert but non-specialist-with-tool versus non-specialist-alone. That study has not been performed, and the figures here do not substitute for it.

## 5. Limitations

### No prospective validation

Every figure in this paper comes from retrospective public data. The system has not been evaluated on images from the deploying clinic’s own equipment, and no prospective performance data existed at the time of writing. This is the single most important limitation and the reason the system is used only with mandatory specialist review. A prospective, patient-level cohort is reserved and withheld from retraining and calibration so that this evaluation, when it is carried out, is independent of the data the model is fitted to.

### Source dependence

Recall varies substantially by source dataset (Table 8). Performance on unfamiliar otoscopes should be assumed closer to the weaker columns than to the pooled figures.

**Table 8.** Recall by source dataset, for conditions present in more than one source.

| **Condition** | **Source A** | **Source B** | **Source C** |
|---|---|---|---|
| Acute otitis media | 85/89 (95.5%) | 9/18 (50.0%) | 13/16 (81.2%) |
| Cerumen impaction | 90/90 (100.0%) | 17/22 (77.3%) | not present |
| Myringosclerosis | 58/91 (63.7%) | 2/4 (50.0%) | not present |
| Normal ear | 88/88 (100.0%) | 67/82 (81.7%) | 40/47 (85.1%) |

### Single-source classes

Eight of twelve classes are drawn from one source each. For these, acquisition style and condition are confounded and cannot be separated within this corpus. The perfect score for chronic otitis media (unspecified) is the clearest instance and should not be quoted as diagnostic performance.

### Small classes

Otitis externa (6 test images), chronic suppurative OM (10), cholesteatoma (9), chronic OM active (13) and *Other findings* (5) have support too small for reliable estimation. Their intervals are wide and are not reported as point estimates to be relied upon. In particular, the absence of missed cholesteatoma is encouraging but rests on 9 test images and does not establish a low miss rate.

### Pooled and overlapping classes

*Other findings* combines foreign bodies, pseudomembranes and ventilation tubes, which share no visual signature and were pooled only because each was too small to learn. The five chronic subtypes overlap in the source labelling, with the unspecified class functioning as their parent.

### Otomycosis is absent

No source contains otomycosis images, so fungal infection cannot be recognised and would be forced into another class. Clinicians can record it only as a free-text diagnosis.

### One image, one ear

Clinical assessment draws on history, examination of both ears, and often audiometry. This system sees a single static image, and its outputs should be weighed accordingly.

### Prevalence dependence

The test split is 69.8% abnormal. Predictive values, particularly PPV, will differ in a screening population with lower prevalence, and the reported 90.2% should not be transferred to such a setting.

### No clinician comparison

The model was not benchmarked against clinicians of any grade, and no study of clinician-plus-model performance has been conducted.

### Label provenance

Ground-truth labels are those supplied by the source datasets. We did not re-adjudicate them, and the accuracy ceiling is therefore set by the quality of the original labelling, which we cannot independently verify. The cross-class review in section 3.2 shows the problem directly: eleven test images have a near-duplicate in the training set under a different label, three of them effectively identical, concentrated in source B. These are inconsistencies in the source collections rather than errors introduced here, but they bound the accuracy any model trained on this corpus can demonstrate.

## 6. Conclusions

A MobileNetV2 classifier with orientation averaging, temperature scaling and a validation-fixed referral threshold achieved 97.0% referral sensitivity and 75.6% specificity on 718 held-out otoscopic images, with AUROC 0.974 for abnormal versus normal. Missed abnormalities were concentrated in myringosclerosis, and no cholesteatoma was missed in this test set.

Two methodological points carry beyond this model. First, near-duplicate control determines the reported score in otoscopic datasets built from video: perceptual hashing was insufficient in our pipeline, and the corrected feature-similarity grouping produced materially lower, and more credible, numbers. Second, performance varies substantially by source dataset even after the framing confound is removed, so single-corpus figures should be read as upper bounds for unfamiliar equipment.

The system is deployed with mandatory ENT review, a scheduled backtest against clinician verdicts, and retraining gated on both a frozen test set and explicit human approval. Its purpose in that deployment is to begin measuring prospective performance on local patients and equipment, which retrospective public data cannot establish. Until that evidence exists, the system is a screening aid under specialist supervision and is not a diagnostic device.

## Data Availability

All data produced in the present study are available upon reasonable request to the authors

## 7. Declarations

### Ethics approval and consent to participate

Ethical approval for this study was obtained from the Nawaloka Hospitals Research and Education Foundation Ethics Review Committee (reference P/26/10/N-0001, approved 9 October 2026). The approval covers the retrospective analysis of public datasets reported here, the prospective collection of otoscopic images and clinician reviews by the deployed system, the use of those reviews for approval-gated retraining, and the clinical governance arrangements set out in section 2.7. The study used only publicly available, de-identified otoscopic image datasets. No patients were recruited, no identifiable patient data were accessed by the investigators, and no intervention was applied to any patient. The deployed system described in the Methods records a hospital record number, age, sex and the ear examined, and stores no name, date of birth or contact detail; it operates under the governance of the Nawaloka Hospitals Research and Education Foundation, and every result it produces is reviewed by a qualified ENT practitioner before it stands as a clinical record.

### Consent for publication

Not applicable. No individually identifiable patient data or images of identifiable persons are included.

### Availability of data and materials

All images were obtained from three openly available otoscopic image datasets published on Kaggle (ucimachinelearning, 2025; Başaran, 2022; Zaka ur Rehman, 2026), with licences given in Table 1, and remain available from their original depositors there. The class assignment applied to those images is our own and is recorded in full in the taxonomy mapping and manifest distributed with the code, so a reader can reconstruct our 12 classes from the original downloads. Reuse of these images is governed by the licences listed in Table 1, and the three differ in ways that bear on redistribution. Source A is released under CC0 1.0 and carries no restriction on reuse. Source C is licensed CC BY-NC-SA 4.0: its NonCommercial term restricts commercial use of material derived from it, and its ShareAlike term would extend to the trained weights if those are treated as an adaptation of the images, so the model weights are not offered under terms less restrictive than that licence and are not available for commercial use. Source B states no licence on its dataset record, so its conditions of reuse are undefined by that record; the only condition its depositor states is that Basaran et al. (2020) be cited, which this paper does, and no further reuse permission has been documented by the depositor. Any party redistributing the images, or weights derived from them, should satisfy itself of these terms. The study manifest, recording for every image its source dataset, class, near-duplicate group and split assignment, is distributed with the code, as are the label ordering, calibration parameters and the machine-generated evaluation report from which every figure in this paper is drawn. Patient images and clinician reviews collected by the deployed system are not available: they are clinical records held by Nawaloka Hospitals and were not used in any analysis reported here.

### Code availability

Source code, trained model weights, the evaluation script that reproduces every reported figure, and the automated test suite are available at https://github.com/GauthamOfficial/OTOMYCOSIS-NHREF.

### Author contributions

Contributions are stated below using the CRediT taxonomy.

- **L. G. Chandrasena:** funding acquisition; resources; supervision.
- **M. T. D. Lakshan:** conceptualisation; supervision; project administration; clinical oversight; writing, review and editing.
- **V. T. K. De Mel:** supervision; investigation; validation; writing, review and editing.
- **B. Kumar Gautham:** conceptualisation; methodology; software; validation; formal analysis; data curation; visualisation; writing, original draft.

The assignment of all 4,763 images to the 12 study classes, described in Methods, was performed by the authors and is credited under data curation.

## Acknowledgements

We thank the depositors of the three public datasets used in this study for making them openly available, and the ENT specialists whose original annotations those datasets carry.

## Use of AI tools

An AI coding assistant (Claude, Anthropic) was used during development of the software and in drafting and editing this manuscript. Every figure reported here was produced by the scripted evaluation of the frozen test split and checked against the machine-generated evaluation artefacts distributed with the code. The authors reviewed all content, verified the reported figures, and take full responsibility for the methods, results and their interpretation. No AI system meets authorship criteria and none is listed as an author, in line with ICMJE guidance.

## Regulatory status

NHREF OtoScan is research software. It is not a registered or certified medical device in any jurisdiction, has not undergone prospective clinical validation, and is deployed only as a screening aid with mandatory review by a qualified ENT practitioner. It must not be used to withhold care. Its intended use was reviewed by the supervising ENT surgeon (M. T. D. Lakshan) and confirmed as research use only; registration with the national medicines regulator would be required before any clinical use beyond research with mandatory specialist review. A prospective, patient-level cohort is reserved for independent performance evaluation: its membership is fixed by the patient’s identifier rather than drawn afresh at each retraining run, and cases belonging to it are excluded from all retraining and from calibration, so that the first prospective estimate of local performance is obtained on patients whose images the model has never been fitted to.

